# Use of Nicotine Products, Prescription Drug Products, and Other Methods to Stop Smoking by US Adults in the 2022 National Health Interview Survey

**DOI:** 10.1101/2024.07.11.24310141

**Authors:** Floe Foxon, Raymond Niaura

## Abstract

Recent data on methods used by adults to stop smoking can inform tobacco control policies. Nationally representative Centers for Disease Control and Prevention survey data from the 2022 National Health Interview Survey (N=27,651) were used to analyze populations of US adults who self-reported having stopped smoking cigarettes for 6 months or longer in the last year and the methods they used, or who did not stop smoking but tried in the last year (n=1,735). In 2022, an estimated 2.9 million [95% CI:2.5 million–3.2 million] US adults had stopped smoking in the past year. Most were male, non-Hispanic White, aged <55 years, college-educated, identified as straight, were not depressed, and currently drank alcohol. The most popular methods used to stop smoking were nicotine products (53.9% [47.4%–60.3%]; 1.5 [1.3– 1.8] million adults), especially e-cigarettes in combination with other methods (40.8% [34.4%– 47.5%]; 1.2 [0.9–1.4] million) and e-cigarettes alone (26.0% [20.4%–32.3%]; 0.7 [0.6–0.9] million). Prescription drug products (8.1% [5.3%–11.8%]; 0.2 [0.1–0.3] million) and non-nicotine, non-prescription drug methods (6.3% (3.9%–9.7%); 0.2 [0.1–0.3] million) were less popular. A further 13.1 [12.2–14.0] million tried but did not stop smoking. Compared to those who tried but didn’t stop smoking, those who successfully stopped were more likely to be younger, degree-educated, and to use e-cigarettes to stop smoking. Many adults still attempt to stop smoking unaided. Interventions to reduce smoking could focus on populations that stopped smoking the least and encourage use of evidence-based methods.

## Introduction

Although considerable progress has been made in the last decade in the US to reduce prevalence of combustible cigarette smoking [1], smoking continues as one of the leading causes of chronic disease [2]. Thus, further reductions in smoking prevalence are crucial to preventing or reducing the effects of chronic disease in the US. A previous study using data from the Population Assessment of Tobacco and Health (PATH) Study found that unaided quit attempts were the most common methods used by US adults to stop smoking from 2013 through 2014 accounting for almost half of quit attempts, with the most frequently used single methods being help from friends and family followed by use of e-cigarettes [3]. Another study found that giving up cigarettes all at once (65.3% of people who smoke and tried to quit), gradually cutting back on cigarettes (62.0%), and substituting some cigarettes with e-cigarettes (35.3%) as the most commonly reported stop-smoking methods during quit attempts from 2014 through 2016 [4]. More recent data are needed to understand who stopped smoking since 2016 and how they did it, particularly with respect to novel nicotine products such as e-cigarettes undergoing FDA review to determine whether or not they may be considered “appropriate for the protection of public health” (e.g. see [5]). The objectives of this study were to analyze demographic populations who self-reported having stopped smoking completely (i.e., self-reported having stopped smoking for ≥6 months in the past year as defined in previous research [6]) or tried to stop smoking, and to quantify the prevalence of the methods they used in the most recent CDC National Health Interview Survey (NHIS) data [7].

## Methods

Data on methods used to stop smoking were sourced from CDC’s NHIS 2022 (total survey sample size N=27,651). These data are nationally representative and have low bias [7]. The 2022 NHIS (fielded January–December 2022) asked respondents who had stopped smoking in the past two years at the time of the survey which methods they used to stop smoking completely. The 2022 survey also asked respondents who were still smoking but had tried to stop in the past one year at the time of the survey which methods they used when they tried. For easier comparison between the ‘stopped smoking completely’ sample (past *two* years at the time of the survey) and the ‘tried’ sample (past *one* year at the time of the survey), we restricted the sample of respondents who stopped smoking in the past two years to just those who stopped smoking in the past one year and who had stopped smoking for 6 months or longer at the time of the survey (i.e., those who had quit for a time 6 months ≤ t ≤ 1 year). Thus, these analyses looked at adults who stopped (N=304) or tried to stop (N=1,431) smoking in the past one year at the time of the 2022 survey. Full details of questionnaire wording are available in online supplemental materials (https://doi.org/10.17605/osf.io/HGMN6).

Although NHIS is an annual survey, other years of NHIS including the 2023 survey and previous recent NHIS questionnaires do not contain questions on methods used to stop smoking, preventing the analysis of trends across time. However, some comparisons can be made to older data in other surveys [3, 4].

The prevalence of each stop-smoking method was calculated with NHIS sampling weights to provide nationally representative population estimates. NHIS stratum and cluster variables were also used to provide 95% CIs. NHIS sampling weights were also used to produce nationally-representative weighted sample sizes, which represent estimates of the corresponding number of US adults at the population level. Demographic characteristics of the sample of adults who stopped smoking or tried to stop (sex, race or ethnicity, age, education, and sexual orientation) were derived from standard NHIS questions, similar to other CDC NHIS publications [8, 9]. Additionally, the mental health status (severity of depressive symptoms - Patient Health Questionnaire depression scale categorization per NHIS definitions) and alcohol drinking status (current, former, or never use per NHIS definitions) of the sample of adults who stopped smoking or tried to stop were also examined.

To determine whether differences between adults who stopped smoking completely and adults who tried to stop smoking but did not stop smoking were significant statistically significant while accounting for the complex survey design, Rao-Scott chi-square tests were used with alpha = 0.05.

Among respondents who tried to stop smoking but did not stop smoking, frequency of smoking was categorized as daily (“now” smoking “every day”) or non-daily (“now” smoking “some days”). The prevalence of each stop-smoking method was calculated for daily and non-daily smoking respondents separately, with Rao-Scott chi-square tests for differences between the two groups.

Only publicly available, de-identified survey data were used; therefore, this study is exempt from NIH human subjects research under NIH exemption 4 and did not require institutional review board review. These analyses were exploratory and were not preregistered.

## Results

### Overall Findings

Overall, an estimated 2.9 million [95% CI: 2.5 million – 3.2 million] US adults had stopped smoking in the past year and had been stopped for 6 months or longer at the time of the 2022 survey (for context, an estimated 29 million adults currently [“every day” or “some days”] smoked cigarettes at the time of the survey).

### Demographic Characteristics

1.6 million [1.3 million – 1.9 million] men and 1.3 million [1.1 million – 1.5 million] women stopped smoking; most were non-Hispanic White adults, most were aged younger than 55 years, most had at least some college education, most self-identified as straight, most had none/minimal severity depressive symptoms, and most currently drank alcohol (Table 1). For reference, current alcohol use prevalence was 69.1% [68.2% – 70.0%] among the general US population in the 2022 NHIS, and 75.0% [72.6% – 77.3%] among adults who stopped or tried to stop smoking. The prevalence of none/minimal severity depressive symptoms was 78.6% [78.0% – 79.3%] among the general US population in the 2022 NHIS and 65.2% [62.6% – 67.7%] among adults who stopped or tried to stop smoking. Compared to those who tried but didn’t stop smoking, those who successfully stopped were more likely to be younger, degree-educated, and gay/lesbian/bisexual, though fewer gay/lesbian/bisexual adults stopped smoking overall compared to straight adults.

**Table 1.** Demographic Characteristics of Adults Who Stopped Smoking (N = 304), or Tried to Stop Smoking but Did Not Stop Smoking (N = 1,431) in the Past Year at the Time of the Survey, National Health Interview Survey 2022^a^.

| Characteristic | Stopped smoking completely<sup>b</sup>, (N = 304)<br>Weighted % (95% CI)<br>Weighted N in millions (95% CI)<br>Unweighted n | Tried to stop smoking but did not stop smoking (N = 1,431)<br>Weighted % (95% CI)<br>Weighted N in millions (95% CI)<br>Unweighted n | Total (stopped smoking completely or tried to stop smoking but did not stop smoking) (N = 1,735)<br>Weighted % (95% CI)<br>Weighted N in millions (95% CI)<br>Unweighted n |
| --- | --- | --- | --- |
| **All adults** | 100<br>2.9 (2.5–3.2)<br>304 | 100<br>13.1 (12.2–14.0)<br>1431 | 100<br>16.0 (15.0–17.0)<br>1735 |
| **Sex** |  |  |  |
| Female | 45.3 (39.1–51.5)<br>1.3 (1.1–1.5)<br>156 | 44.2 (41.1–47.2)<br>5.8 (5.3–6.3)<br>687 | 44.4 (41.6–47.1)<br>7.1 (6.5–7.7)<br>843 |
| Male | 54.7 (48.5–60.9)<br>1.6 (1.3–1.9)<br>148 | 55.8 (52.8–58.9)<br>7.3 (6.7–7.9)<br>744 | 55.6 (52.9–58.4)<br>8.9 (8.2–9.6)<br>892 |
| **Race or ethnicity** |  |  |  |
| Hispanic | 15.2 (10.8–20.6)<br>0.4 (0.3–0.6)<br>43 | 12.0 (9.9–14.4)<br>1.6 (1.3–1.9)<br>152 | 12.6 (10.6–14.9)<br>2.0 (1.7–2.4)<br>195 |
| Non-Hispanic Black | 11.2 (7.4–16.1)<br>0.3 (0.2–0.5)<br>35 | 16.1 (13.7–18.6)<br>2.1 (1.8–2.4)<br>247 | 15.2 (13.1–17.5)<br>2.4 (2.1–2.8)<br>282 |
| Non-Hispanic White | 67.3 (60.9–73.2)<br>1.9 (1.6–2.2)<br>207 | 65.1 (62.1–68.0)<br>8.5 (7.9–9.2)<br>937 | 65.5 (62.7–68.1)<br>10.5 (9.7–11.2)<br>1144 |
| Non-Hispanic Other | 6.3 (3.5–10.3)<br>0.2 (0.1–0.3)<br>19 | 6.8 (5.4–8.6)<br>0.9 (0.7–1.1)<br>95 | 6.7 (5.4–8.3)<br>1.1 (0.8–1.3)<br>114 |
| **Age (Years)\*** |  |  |  |
| 18–34 | 41.9 (35.3–48.8)<br>1.2 (1.0–1.4)<br>111 | 24.7 (21.9–27.6)<br>3.2 (2.8–3.7)<br>268 | 27.8 (25.2–30.5)<br>4.4 (3.9–5.0)<br>379 |
| 35–54 | 34.2 (27.9–40.9)<br>1.0 (0.8–1.2)<br>102 | 39.6 (36.4–42.8)<br>5.2 (4.7–5.7)<br>512 | 38.6 (35.8–41.4)<br>6.2 (5.6–6.8)<br>614 |
| 55 or older | 23.9 (18.8–29.6)<br>0.7 (0.5–0.9)<br>91 | 35.7 (33.0–38.6)<br>4.7 (4.3–5.1)<br>651 | 33.6 (31.1–36.2)<br>5.4 (4.9–5.9)<br>742 |
| **Education\*** |  |  |  |
| ≤12 years (no diploma) | 9.4 (5.9–14.1)<br>0.3 (0.1–0.4)<br>27 | 18.5 (16.0–21.3)<br>2.4 (2.0–2.8)<br>220 | 16.9 (14.7–19.3)<br>2.7 (2.3–3.1)<br>247 |
| GED certificate or high school diploma | 34.4 (28.5–40.7)<br>1.0 (0.8–1.2)<br>95 | 38.5 (35.5–41.7)<br>5.0 (4.5–5.5)<br>519 | 37.8 (35.0–40.7)<br>6.0 (5.4–6.5)<br>614 |
| Some college (no degree) | 18.8 (14.4–23.9)<br>0.5 (0.4–0.7)<br>61 | 17.1 (15.1–19.3)<br>2.2 (1.9–2.5)<br>258 | 17.4 (15.6–19.4)<br>2.8 (2.4–3.1)<br>319 |
| Associate, undergraduate, or graduate degree | 37.3 (31.1–43.9)<br>1.1 (0.8–1.3)<br>117 | 25.8 (23.2–28.5)<br>3.4 (2.9–3.8)<br>426 | 27.9 (25.4–30.4)<br>4.4 (4.0–4.9)<br>543 |
| **Sexual orientation\*** |  |  |  |
| Gay/lesbian/bisexual | 12.8 (8.7–17.9)<br>0.4 (0.2–0.5)<br>37 | 5.8 (4.5–7.4)<br>0.8 (0.6–1.0)<br>82 | 7.1 (5.7–8.6)<br>1.1 (0.9–1.4)<br>119 |
| Straight | 82.8 (76.8–87.8)<br>2.4 (2.0–2.7)<br>254 | 90.0 (88.1–91.7)<br>11.8 (11.0–12.6)<br>1293 | 88.7 (86.9–90.4)<br>14.2 (13.3–15.1)<br>1547 |
| Other | 4.4 (1.8–8.7)<br>0.1 (0.0–0.2)<br>13 | 4.1 (3.0–5.6)<br>0.5 (0.4–0.7)<br>56 | 4.2 (3.1–5.5)<br>0.7 (0.5–0.9)<br>69 |
| **Severity of depressive symptoms - Patient Health Questionnaire depression scale categorization** |  |  |  |
| None/Minimal | 63.7 (57.1–70.1)<br>1.8 (1.5–2.1)<br>191 | 65.5 (62.6–68.3)<br>8.6 (7.9–9.3)<br>922 | 65.2 (62.6–67.7)<br>10.4 (9.6–11.2)<br>1113 |
| Mild | 20.7 (15.8–26.4)<br>0.6 (0.4–0.7)<br>64 | 19.5 (17.2–22.0)<br>2.6 (2.2–2.9)<br>278 | 19.7 (17.6–21.9)<br>3.1 (2.8–3.5)<br>342 |
| Moderate | 7.7 (4.8–11.5)<br>0.2 (0.1–0.3)<br>27 | 8.3 (6.8–10.0)<br>1.1 (0.9–1.3)<br>128 | 8.2 (6.8–9.7)<br>1.3 (1.1–1.5)<br>155 |
| Severe | 7.9 (4.7–12.3)<br>0.2 (0.1–0.3)<br>20 | 6.7 (5.2–8.5)<br>0.9 (0.7–1.1)<br>100 | 6.9 (5.5–8.6)<br>1.1 (0.9–1.3)<br>120 |
| **Alcohol drinking status** |  |  |  |
| Current | 80.1 (74.5–84.9)<br>2.3 (1.9–2.6)<br>231 | 73.9 (71.2–76.5)<br>9.5 (8.7–10.2)<br>1000 | 75.0 (72.6–77.3)<br>11.7 (10.9–12.6)<br>1231 |
| Former | 16.6 (12.2–21.8)<br>0.5 (0.3–0.6)<br>58 | 21.0 (18.6–23.6)<br>2.7 (2.4–3.0)<br>332 | 20.2 (18.1–22.5)<br>3.2 (2.8–3.5)<br>390 |
| Never | 3.3 (1.4–6.6)<br>0.1 (0.0–0.2)<br>10 | 5.1 (3.8–6.6)<br>0.6 (0.5–0.8)<br>72 | 4.7 (3.6–6.1)<br>0.7 (0.5–0.9)<br>82 |

\* Indicates a statistically significant difference between adults who stopped smoking completely and adults who tried to stop smoking but did not stop smoking (Rao-Scott chi-square test; $p < 0.05$ ).
<sup>a</sup> Following Cornelius et al. [8], modified Clopper-Pearson confidence limits for percentages were calculated by using the Korn and Graubard method.
<sup>b</sup> Reported having stopped smoking for 6 months or longer in the past year, as defined in previous research [6].

### Stop-Smoking Methods

Among those who stopped smoking, the most commonly reported listed methods were nicotine products (53.9%, 1.5 million US adults), primarily e-cigarettes used alone or in combination with other methods (40.8%, 1.2 million US adults; Table 2). Among those who stopped smoking using e-cigarettes, 69.3% currently used e-cigarettes (i.e. “now” used e-cigarettes “every day” or “some days”), and the remaining 30.7% formerly used e-cigarettes (i.e. “now” used e-cigarettes “not at all”). The least commonly reported listed methods were non-nicotine, non-prescription drug methods (including a quit line, counseling or clinic, class, or group; 6.3%, 0.2 million US adults). Of the listed methods, the most commonly reported exclusive method selected was e-cigarettes; 26.0% (0.7 million US adults) of adults who stopped smoking for 6 months or longer at the time of the 2022 survey selected e-cigarettes as their only listed method.

**Table 2.**
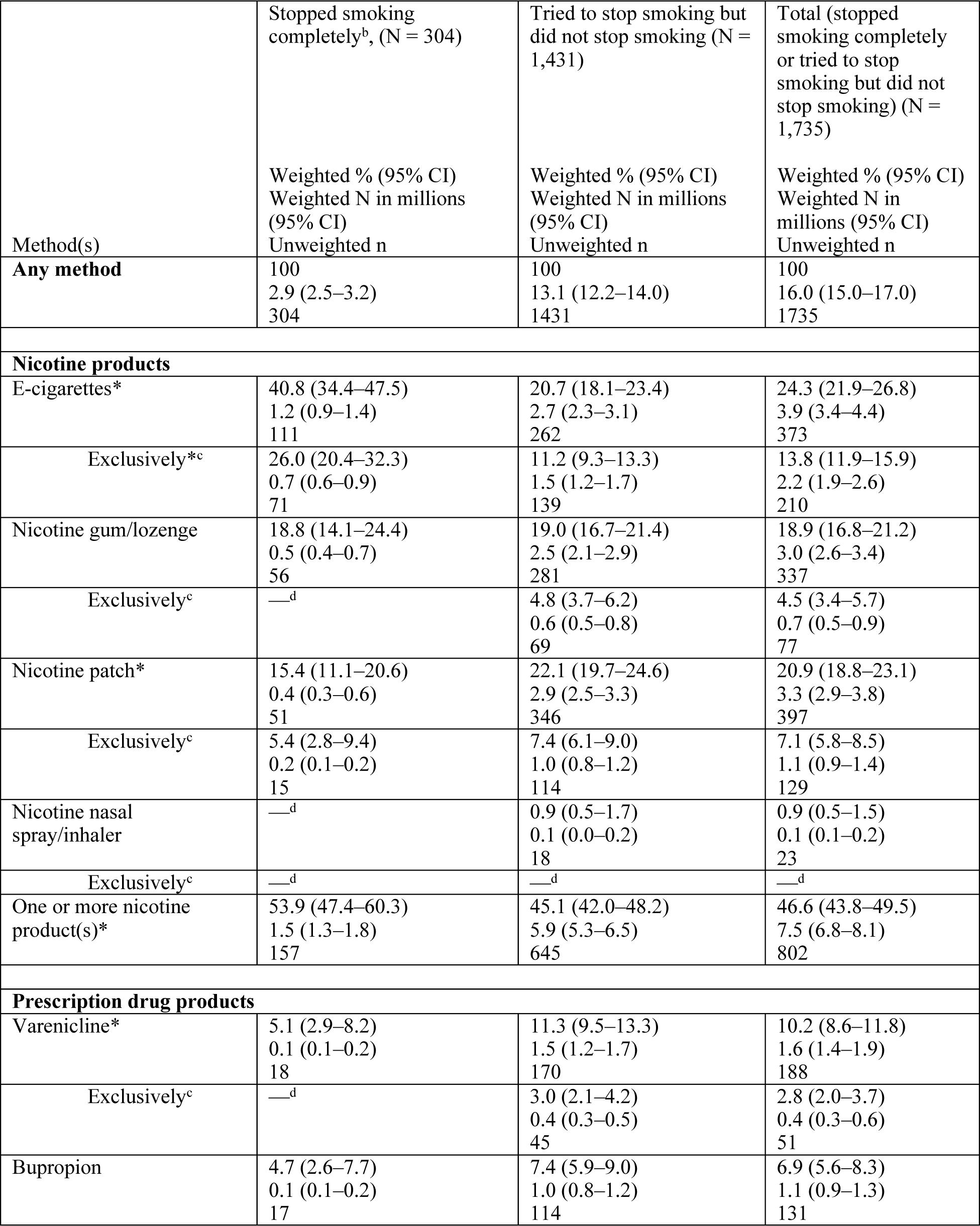
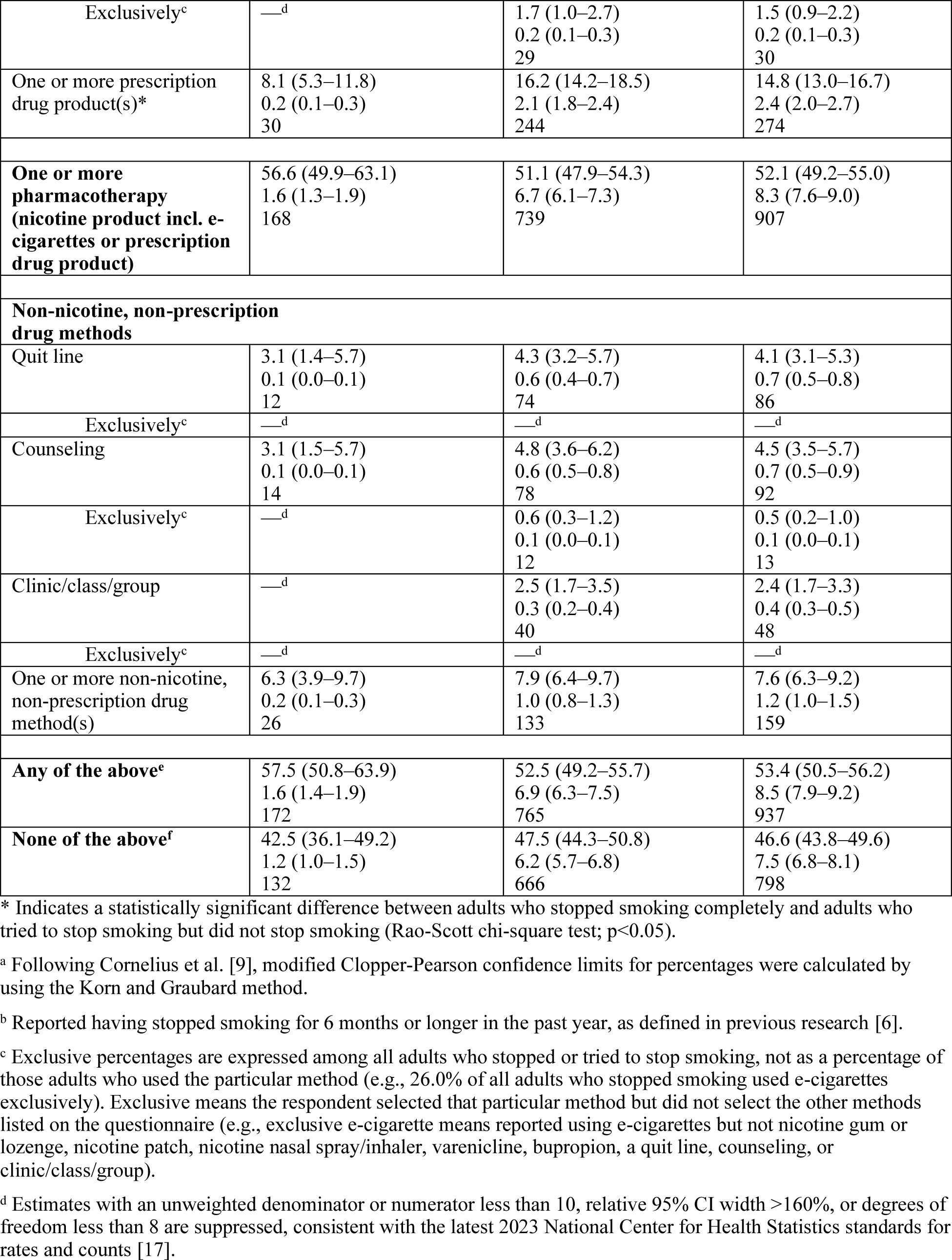
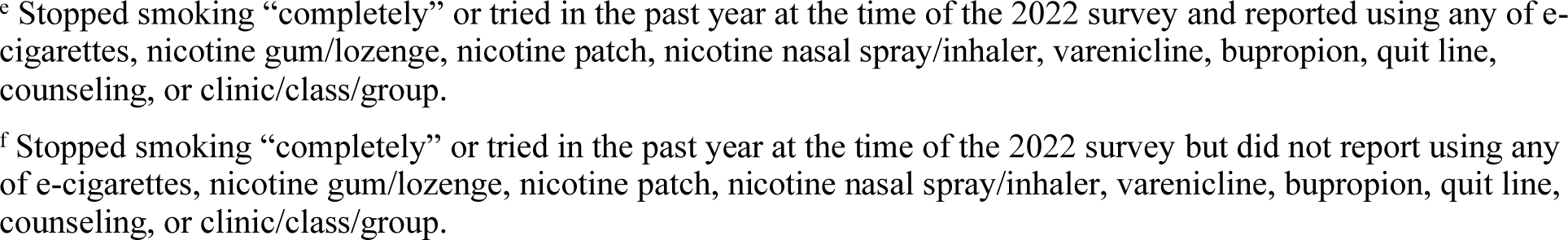
Methods Used by Adults Who Stopped Smoking (N = 304), or Tried to Stop Smoking but Did Not Stop Smoking (N = 1,431) in the Past Year at the Time of the Survey, National Health Interview Survey, 2022^a^.

| Method(s) | Stopped smoking completely<sup>b</sup>, (N = 304)<br>Weighted % (95% CI)<br>Weighted N in millions (95% CI)<br>Unweighted n | Tried to stop smoking but did not stop smoking (N = 1,431)<br>Weighted % (95% CI)<br>Weighted N in millions (95% CI)<br>Unweighted n | Total (stopped smoking completely or tried to stop smoking but did not stop smoking) (N = 1,735)<br>Weighted % (95% CI)<br>Weighted N in millions (95% CI)<br>Unweighted n |
| --- | --- | --- | --- |
| **Any method** | 100<br>2.9 (2.5–3.2)<br>304 | 100<br>13.1 (12.2–14.0)<br>1431 | 100<br>16.0 (15.0–17.0)<br>1735 |
| **Nicotine products** |  |  |  |
| E-cigarettes* | 40.8 (34.4–47.5)<br>1.2 (0.9–1.4)<br>111 | 20.7 (18.1–23.4)<br>2.7 (2.3–3.1)<br>262 | 24.3 (21.9–26.8)<br>3.9 (3.4–4.4)<br>373 |
| Exclusively*<sup>c</sup> | 26.0 (20.4–32.3)<br>0.7 (0.6–0.9)<br>71 | 11.2 (9.3–13.3)<br>1.5 (1.2–1.7)<br>139 | 13.8 (11.9–15.9)<br>2.2 (1.9–2.6)<br>210 |
| Nicotine gum/lozenge | 18.8 (14.1–24.4)<br>0.5 (0.4–0.7)<br>56 | 19.0 (16.7–21.4)<br>2.5 (2.1–2.9)<br>281 | 18.9 (16.8–21.2)<br>3.0 (2.6–3.4)<br>337 |
| Exclusively<sup>c</sup> | —<sup>d</sup> | 4.8 (3.7–6.2)<br>0.6 (0.5–0.8)<br>69 | 4.5 (3.4–5.7)<br>0.7 (0.5–0.9)<br>77 |
| Nicotine patch* | 15.4 (11.1–20.6)<br>0.4 (0.3–0.6)<br>51 | 22.1 (19.7–24.6)<br>2.9 (2.5–3.3)<br>346 | 20.9 (18.8–23.1)<br>3.3 (2.9–3.8)<br>397 |
| Exclusively<sup>c</sup> | 5.4 (2.8–9.4)<br>0.2 (0.1–0.2)<br>15 | 7.4 (6.1–9.0)<br>1.0 (0.8–1.2)<br>114 | 7.1 (5.8–8.5)<br>1.1 (0.9–1.4)<br>129 |
| Nicotine nasal spray/inhaler | —<sup>d</sup> | 0.9 (0.5–1.7)<br>0.1 (0.0–0.2)<br>18 | 0.9 (0.5–1.5)<br>0.1 (0.1–0.2)<br>23 |
| Exclusively<sup>c</sup> | —<sup>d</sup> | —<sup>d</sup> | —<sup>d</sup> |
| One or more nicotine product(s)* | 53.9 (47.4–60.3)<br>1.5 (1.3–1.8)<br>157 | 45.1 (42.0–48.2)<br>5.9 (5.3–6.5)<br>645 | 46.6 (43.8–49.5)<br>7.5 (6.8–8.1)<br>802 |
| **Prescription drug products** |  |  |  |
| Varenicline* | 5.1 (2.9–8.2)<br>0.1 (0.1–0.2)<br>18 | 11.3 (9.5–13.3)<br>1.5 (1.2–1.7)<br>170 | 10.2 (8.6–11.8)<br>1.6 (1.4–1.9)<br>188 |
| Exclusively<sup>c</sup> | —<sup>d</sup> | 3.0 (2.1–4.2)<br>0.4 (0.3–0.5)<br>45 | 2.8 (2.0–3.7)<br>0.4 (0.3–0.6)<br>51 |
| Bupropion | 4.7 (2.6–7.7)<br>0.1 (0.1–0.2)<br>17 | 7.4 (5.9–9.0)<br>1.0 (0.8–1.2)<br>114 | 6.9 (5.6–8.3)<br>1.1 (0.9–1.3)<br>131 |
| Exclusively<sup>c</sup> | —<sup>d</sup> | 1.7 (1.0–2.7)<br>0.2 (0.1–0.3)<br>29 | 1.5 (0.9–2.2)<br>0.2 (0.1–0.3)<br>30 |
| One or more prescription drug product(s)* | 8.1 (5.3–11.8)<br>0.2 (0.1–0.3)<br>30 | 16.2 (14.2–18.5)<br>2.1 (1.8–2.4)<br>244 | 14.8 (13.0–16.7)<br>2.4 (2.0–2.7)<br>274 |
| **One or more pharmacotherapy (nicotine product incl. e-cigarettes or prescription drug product)** | 56.6 (49.9–63.1)<br>1.6 (1.3–1.9)<br>168 | 51.1 (47.9–54.3)<br>6.7 (6.1–7.3)<br>739 | 52.1 (49.2–55.0)<br>8.3 (7.6–9.0)<br>907 |
| **Non-nicotine, non-prescription drug methods** |  |  |  |
| Quit line | 3.1 (1.4–5.7)<br>0.1 (0.0–0.1)<br>12 | 4.3 (3.2–5.7)<br>0.6 (0.4–0.7)<br>74 | 4.1 (3.1–5.3)<br>0.7 (0.5–0.8)<br>86 |
| Exclusively<sup>c</sup> | —<sup>d</sup> | —<sup>d</sup> | —<sup>d</sup> |
| Counseling | 3.1 (1.5–5.7)<br>0.1 (0.0–0.1)<br>14 | 4.8 (3.6–6.2)<br>0.6 (0.5–0.8)<br>78 | 4.5 (3.5–5.7)<br>0.7 (0.5–0.9)<br>92 |
| Exclusively<sup>c</sup> | —<sup>d</sup> | 0.6 (0.3–1.2)<br>0.1 (0.0–0.1)<br>12 | 0.5 (0.2–1.0)<br>0.1 (0.0–0.1)<br>13 |
| Clinic/class/group | —<sup>d</sup> | 2.5 (1.7–3.5)<br>0.3 (0.2–0.4)<br>40 | 2.4 (1.7–3.3)<br>0.4 (0.3–0.5)<br>48 |
| Exclusively<sup>c</sup> | —<sup>d</sup> | —<sup>d</sup> | —<sup>d</sup> |
| One or more non-nicotine, non-prescription drug method(s) | 6.3 (3.9–9.7)<br>0.2 (0.1–0.3)<br>26 | 7.9 (6.4–9.7)<br>1.0 (0.8–1.3)<br>133 | 7.6 (6.3–9.2)<br>1.2 (1.0–1.5)<br>159 |
| **Any of the above<sup>e</sup>** | 57.5 (50.8–63.9)<br>1.6 (1.4–1.9)<br>172 | 52.5 (49.2–55.7)<br>6.9 (6.3–7.5)<br>765 | 53.4 (50.5–56.2)<br>8.5 (7.9–9.2)<br>937 |
| **None of the above<sup>f</sup>** | 42.5 (36.1–49.2)<br>1.2 (1.0–1.5)<br>132 | 47.5 (44.3–50.8)<br>6.2 (5.7–6.8)<br>666 | 46.6 (43.8–49.6)<br>7.5 (6.8–8.1)<br>798 |

\* Indicates a statistically significant difference between adults who stopped smoking completely and adults who tried to stop smoking but did not stop smoking (Rao-Scott chi-square test; $p < 0.05$ ).
<sup>a</sup> Following Cornelius et al. [9], modified Clopper-Pearson confidence limits for percentages were calculated by using the Korn and Graubard method.
<sup>b</sup> Reported having stopped smoking for 6 months or longer in the past year, as defined in previous research [6].
<sup>c</sup> Exclusive percentages are expressed among all adults who stopped or tried to stop smoking, not as a percentage of those adults who used the particular method (e.g., 26.0% of all adults who stopped smoking used e-cigarettes exclusively). Exclusive means the respondent selected that particular method but did not select the other methods listed on the questionnaire (e.g., exclusive e-cigarette means reported using e-cigarettes but not nicotine gum or lozenge, nicotine patch, nicotine nasal spray/inhaler, varenicline, bupropion, a quit line, counseling, or clinic/class/group).
<sup>d</sup> Estimates with an unweighted denominator or numerator less than 10, relative 95% CI width >160%, or degrees of freedom less than 8 are suppressed, consistent with the latest 2023 National Center for Health Statistics standards for rates and counts [17].
<sup>e</sup> Stopped smoking “completely” or tried in the past year at the time of the 2022 survey and reported using any of e-cigarettes, nicotine gum/lozenge, nicotine patch, nicotine nasal spray/inhaler, varenicline, bupropion, quit line, counseling, or clinic/class/group.
<sup>f</sup> Stopped smoking “completely” or tried in the past year at the time of the 2022 survey but did not report using any of e-cigarettes, nicotine gum/lozenge, nicotine patch, nicotine nasal spray/inhaler, varenicline, bupropion, quit line, counseling, or clinic/class/group.

**Table 3.** Methods Used by Adults Who Tried to Stop Smoking but Did Not Stop Smoking (N = 1,431) in the Past Year at the Time of the Survey by Smoking Frequency, National Health Interview Survey, 2022^a^.

| Method(s) | Daily Smoking<sup>b</sup>, (N = 990)<br>Weighted % (95% CI)<br>Weighted N in millions (95% CI)<br>Unweighted n | Non-Daily Smoking<sup>b</sup> (N = 441)<br>Weighted % (95% CI)<br>Weighted N in millions (95% CI)<br>Unweighted n |
| --- | --- | --- |
| **Any method** | 100<br>8.9 (8.2–9.6)<br>990 | 100<br>4.2 (3.7–4.7)<br>441 |
| **Nicotine products** |  |  |
| E-cigarettes | 19.7 (16.6–23.1)<br>1.8 (1.4–2.1)<br>180 | 22.8 (18.0–28.2)<br>1.0 (0.7–1.2)<br>82 |
| Exclusively<sup>*c</sup> | 9.5 (7.3–12.1)<br>0.8 (0.6–1.1)<br>87 | 14.6 (10.7–19.4)<br>0.6 (0.4–0.8)<br>52 |
| Nicotine gum/lozenge<sup>*</sup> | 21.5 (18.6–24.6)<br>1.9 (1.6–2.2)<br>222 | 13.7 (10.3–17.7)<br>0.6 (0.4–0.7)<br>59 |
| Exclusively<sup>c</sup> | 4.8 (3.5–6.4)<br>0.4 (0.3–0.6)<br>52 | 4.8 (2.6–8.1)<br>0.2 (0.1–0.3)<br>17 |
| Nicotine patch<sup>*</sup> | 25.8 (22.8–28.9)<br>2.3 (2.0–2.6)<br>272 | 14.4 (10.9–18.5)<br>0.6 (0.4–0.8)<br>74 |
| Exclusively<sup>*c</sup> | 9.1 (7.3–11.3)<br>0.8 (0.6–1.0)<br>91 | 3.8 (2.2–6.1)<br>0.2 (0.1–0.2)<br>23 |
| Nicotine nasal spray/inhaler | —<sup>d</sup> | —<sup>d</sup> |
| Exclusively<sup>c</sup> | —<sup>d</sup> | —<sup>d</sup> |
| One or more nicotine product(s)<sup>*</sup> | 47.8 (44.0–51.6)<br>4.3 (3.8–4.8)<br>482 | 39.3 (33.7–45.2)<br>1.7 (1.3–2.0)<br>163 |
| **Prescription drug products** |  |  |
| Varenicline<sup>*</sup> | 12.8 (10.4–15.4)<br>1.1 (0.9–1.4)<br>131 | 8.1 (5.5–11.4)<br>0.3 (0.2–0.5)<br>39 |
| Exclusively<sup>c</sup> | 3.3 (2.1–4.8)<br>0.3 (0.2–0.4)<br>32 | 2.6 (1.3–4.7)<br>0.1 (0.0–0.2)<br>13 |
| Bupropion<sup>*</sup> | 8.7 (6.8–11.0)<br>0.8 (0.6–1.0)<br>90 | 4.4 (2.7–6.9)<br>0.2 (0.1–0.3)<br>24 |
| Exclusively<sup>c</sup> | 2.0 (1.1–3.3)<br>0.2 (0.1–0.3)<br>20 | —<sup>d</sup> |
| One or more prescription drug product(s)<sup>*</sup> | 18.5 (15.8–21.5)<br>1.7 (1.4–1.9)<br>188 | 11.4 (8.4–15.0)<br>0.5 (0.3–0.6)<br>56 |
| **One or more pharmacotherapy (nicotine product incl. e-cigarettes or prescription drug product)\*** | 54.9 (50.9–58.7)<br>4.9 (4.4–5.4)<br>554 | 43.2 (37.4–49.1)<br>1.8 (1.5–2.1)<br>185 |
| **Non-nicotine, non-prescription drug methods** |  |  |
| Quit line* | 5.2 (3.7–7.0)<br>0.5 (0.3–0.6)<br>61 | 2.6 (1.2–4.7)<br>0.1 (0.0–0.2)<br>13 |
| Exclusively<sup>c</sup> | —<sup>d</sup> | —<sup>d</sup> |
| Counseling* | 5.6 (4.1–7.4)<br>0.5 (0.4–0.6)<br>63 | 3.0 (1.5–5.3)<br>0.1 (0.0–0.2)<br>15 |
| Exclusively<sup>c</sup> | —<sup>d</sup> | —<sup>d</sup> |
| Clinic/class/group | 2.8 (1.8–4.1)<br>0.2 (0.1–0.3)<br>32 | —<sup>d</sup> |
| Exclusively<sup>c</sup> | —<sup>d</sup> | —<sup>d</sup> |
| One or more non-nicotine, non-prescription drug method(s)* | 9.4 (7.4–11.8)<br>0.8 (0.6–1.0)<br>108 | 4.8 (2.9–7.3)<br>0.2 (0.1–0.3)<br>25 |
| **Any of the above\*<sup>e</sup>** | 56.5 (52.5–60.4)<br>5.0 (4.5–5.6)<br>575 | 44.0 (38.2–49.9)<br>1.9 (1.5–2.2)<br>190 |
| **None of the above\*<sup>f</sup>** | 43.5 (39.6–47.5)<br>3.9 (3.4–4.3)<br>415 | 56.0 (50.1–61.8)<br>2.4 (2.0–2.7)<br>251 |

\* Indicates a statistically significant difference between adults who smoked daily and adults who smoked non-daily (Rao-Scott chi-square test; $p < 0.05$ ).
<sup>a</sup> Following Cornelius et al. [9], modified Clopper-Pearson confidence limits for percentages were calculated by using the Korn and Graubard method.
<sup>b</sup> Daily smoking is defined as having ever smoked 100 cigarettes (lifetime) and “now” smoking cigarettes “every day”. Non-daily smoking is defined as having ever smoked 100 cigarettes (lifetime) and “now” smoking cigarettes “some days”.
<sup>c</sup> Exclusive percentages are expressed among all adults who tried to stop smoking, not as a percentage of those adults who used the particular method. Exclusive means the respondent selected that particular method but did not select the other methods listed on the questionnaire (e.g., exclusive e-cigarette means reported using e-cigarettes but not nicotine gum or lozenge, nicotine patch, nicotine nasal spray/inhaler, varenicline, bupropion, a quit line, counseling, or clinic/class/group).
<sup>d</sup> Estimates with an unweighted denominator or numerator less than 10, relative 95% CI width >160%, or degrees of freedom less than 8 are suppressed, consistent with the latest 2023 National Center for Health Statistics standards for rates and counts [17].
<sup>e</sup> Tried to stop smoking in the past year at the time of the 2022 survey and reported using any of e-cigarettes, nicotine gum/lozenge, nicotine patch, nicotine nasal spray/inhaler, varenicline, bupropion, quit line, counseling, or clinic/class/group.
<sup>f</sup> Tried to stop smoking in the past year at the time of the 2022 survey but did not report using any of e-cigarettes, nicotine gum/lozenge, nicotine patch, nicotine nasal spray/inhaler, varenicline, bupropion, quit line, counseling, or clinic/class/group.

In the same period, 13.1 million [12.2 million – 14.0 million] tried to stop but were unsuccessful. Compared to those who tried but did not stop smoking, those who successfully stopped were more likely to report using e-cigarettes to stop smoking; and less likely to report using nicotine patches and prescription drugs to stop smoking.

Among those who tried but did not stop smoking, compared to those who smoked non-daily, those who smoked daily were more likely to have used one or more pharmacotherapy (nicotine products and prescription drugs) and non-nicotine, non-prescription drug methods in their attempt to stop smoking. I.e., compared to those who smoked daily, those who smoked non-daily were more likely to not have tried any of the surveyed methods when they attempted to stop smoking.

## Discussion

This study analyzed populations of US adults in NHIS 2022 who stopped or tried to stop smoking and the methods they used. Approximately 29 million adults currently smoked cigarettes in 2022. The number of adults who stopped smoking completely was only one tenth of that number, and less than half of those 29 million who remained smoking tried to stop smoking (but did not stop). These findings suggest that more efforts are required to encourage adults who smoke to try to stop smoking in the first place.

46.6% of adults (including 42.5% of adults who stopped smoking and 47.5% of adults who tried but did not stop smoking) did not report using any of the surveyed evidence-based methods to stop smoking. This finding is not dissimilar to the finding in the PATH 2013–2014 data by Rodu and Plurphanswat [3], who reported that 33.5% of adults who currently smoked (and who tried to stop) and 32.8% of adults who formerly smoked reported having attempted to stop smoking completely unaided. Methods containing nicotine, primarily e-cigarettes, were the most commonly-reported methods that were explicitly surveyed. Caraballo et al. [4] found that in 2014–2016 data, switching to e-cigarettes was reported by 35.3% of US adults who smoked cigarettes in their most recent attempt, while Rodu and Plurphanswat [3] found that in 2013– 2014 data, e-cigarettes were the second most frequently used single method to stop smoking (32% of all single methods). In comparison, the present study found in 2022 data that e-cigarettes were used to stop smoking by 40.8% of adults who stopped smoking and 20.7% of adults who tried but did not stop smoking. While noting the necessity of preventing access to nicotine products by nonsmoking populations such as adolescents, the high prevalence of e-cigarette use to stop smoking in this study may provide support for FDA’s “nicotine-focused framework for public health” [10], which describes noncombustible nicotine products as “a promising foundation for a comprehensive approach to tobacco harm reduction.” In recent years, FDA have authorized a number of e-cigarette products as “appropriate for the protection of public health” [5]. CDC also state that “[e]-cigarettes have the potential to benefit adults who smoke . . . if used as a complete substitute for regular cigarettes” [11].

The least commonly-reported methods to stop smoking were the non-nicotine, non-prescription drug methods, including quit lines, counselling, and clinics/classes/groups. These methods were reported by just 6.3% of adults who stopped smoking and 7.9% of adults who tried but did not stop smoking. This finding is unsurprising since randomized controlled trials have consistently shown pharmacological interventions - including e-cigarettes - to be superior to non-pharmacological controls in terms of smoking abstinence efficacy [12]. While non-pharmacological methods clearly work for some adults and should not be discouraged, given the urgency of the need to reduce morbidity and mortality from cigarette smoking, it may be preferable from a public health perspective to encourage adults who smoke to try the most efficacious stop-smoking methods first.

Compared to those who tried but didn’t stop smoking, those who successfully stopped were more likely to be younger, degree-educated, and gay/lesbian/bisexual, though fewer gay/lesbian/bisexual adults stopped smoking compared to straight adults. This finding highlights disparities in smoking cessation; it is important to ensure that specific demographic populations do not get left behind in efforts to curb smoking. Additionally, those who successfully stopped smoking were more likely to have used e-cigarettes to stop smoking, and were less likely to have used nicotine patches and prescription drugs to stop smoking. It is possible that these differences are mediated by demographic differences (though model-based comparisons may not be appropriate for these data due to low sample size) and by barriers to access (e.g. out-of-pocket pharmacy costs [13] and other access restrictions [14]).

This study’s strengths include use of a large, nationally representative survey with low bias administered by CDC. NHIS results are thought to be generalizable to the entire US population, because that is the purpose of its sample design by CDC’s National Center for Health Statistics [7, 15]. Another strength is the direct measures of stopping smoking (i.e., survey questions answered by adults who actually stopped smoking) as opposed to indirect measures of stopping smoking (e.g., models).

This study also has limitations. First is the inability to parse out individual methods in the “none of the above” category, which is an unavoidable limitation of the NHIS questionnaire design. Because “none of the above” was inferred from respondents who stopped or tried but who did not select any of the surveyed items, the “none of the above” category may be underrepresented in these analyses. Furthermore, the questionnaire did not ask respondents about the duration of treatment use. Future questionnaires should ask respondents for these details. Recall bias may also limit the accuracy of these findings [16]. Additionally, we chose to treat respondents who responded with “don’t know” as missing, which has its limitations. Also importantly, the NHIS 2022 data did not explicitly survey some of the other more recent stop-smoking methods such as nicotine pouches, snus, heat-not-burn or heated tobacco products, and others – these should also be surveyed in future questionnaires.

In conclusion, more younger adults are stopping smoking compared to older adults, which may leave the latter at risk of being ‘left behind’ in efforts to stop smoking. Nicotine-containing methods, especially e-cigarettes, are by far the most commonly-reported methods used to stop smoking among the evidence-based methods surveyed in NHIS. Adults who stopped smoking completely are more likely to report using e-cigarettes than those who tried but did not stop smoking. These findings may support the basis for harm reduction in tobacco control. However, many adults still attempt to stop smoking without evidence-based methods. Future efforts to reduce smoking among US adults should focus on the demographic populations that stopped smoking the least and encourage use of evidence-based methods.

## Data Availability

Data are available at: https://www.cdc.gov/nchs/nhis/index.htm

## Acknowledgements

An early version of this research was presented in poster form at the 2024 Society for Research on Nicotine and Tobacco (SRNT) annual meeting (https://doi.org/10.31219/osf.io/2yuw4). The authors thank anonymous reviewers and participants at the 2024 SRNT meeting for helpful comments and suggestions. F.F. thanks Mark Sembower of Pinney Associates for discussion. This work received no financial support.

## Statement of Data Availability

NHIS data are publicly available at https://www.cdc.gov/nchs/nhis/index.htm

